# A Multicenter Evaluation of the Impact of Therapies on Deep Learning-based Electrocardiographic Hypertrophic Cardiomyopathy Markers

**DOI:** 10.1101/2024.01.15.24301011

**Authors:** Lovedeep S Dhingra, Veer Sangha, Arya Aminorroaya, Robyn Bryde, Andrew Gaballa, Adel H Ali, Nandini Mehra, Harlan M. Krumholz, Sounok Sen, Christopher M Kramer, Matthew W Martinez, Milind Y Desai, Evangelos K Oikonomou, Rohan Khera

## Abstract

**Background:** Artificial intelligence-enhanced electrocardiography (AI-ECG) can identify hypertrophic cardiomyopathy (HCM) on 12-lead ECGs and offers a novel way to monitor treatment response. While the surgical or percutaneous reduction of the interventricular septum (SRT) represented initial HCM therapies, mavacamten offers an oral alternative.

**Objective:** To evaluate biological response to SRT and mavacamten.

**Methods:** We applied an AI-ECG model for HCM detection to ECG images from patients who underwent SRT across three sites: Yale New Haven Health System (YNHHS), Cleveland Clinic Foundation (CCF), and Atlantic Health System (AHS); and to ECG images from patients receiving mavacamten at YNHHS.

**Results:** A total of 70 patients underwent SRT at YNHHS, 100 at CCF, and 145 at AHS. At YNHHS, there was no significant change in the AI-ECG HCM score before versus after SRT (pre-SRT: median 0.55 [IQR 0.24–0.77] vs post-SRT: 0.59 [0.40–0.75]). The AI-ECG HCM scores also did not improve post SRT at CCF (0.61 [0.32–0.79] vs 0.69 [0.52–0.79]) and AHS (0.52 [0.35–0.69] vs 0.61 [0.49–0.70]). Among 36 YNHHS patients on mavacamten therapy, the median AI-ECG score before starting mavacamten was 0.41 (0.22–0.77), which decreased significantly to 0.28 (0.11–0.50, *p* <0.001 by Wilcoxon signed-rank test) at the end of a median follow-up period of 237 days.

**Conclusions:** The lack of improvement in AI-based HCM score with SRT, in contrast to a significant decrease with mavacamten, suggests the potential role of AI-ECG for serial monitoring of pathophysiological improvement in HCM at the point-of-care using ECG images.

## INTRODUCTION

Artificial intelligence-enhanced electrocardiography (AI-ECG) interpretation can identify hypertrophic cardiomyopathy (HCM) on 12-lead ECGs and represents a novel way to monitor response to treatment.^1–4^ HCM is a genetically determined disease that affects up to 1 in every 200 people globally and is among the leading causes of sudden cardiac death.^5, 6^ Among patients with HCM, mavacamten is the first cardiac myosin inhibitor to have shown efficacy in relieving hemodynamic obstruction and improving health status.^7–9^ Before the emergence of mavacamten, septal reduction therapy (SRT), which includes alcohol septal ablation and ventricular myectomy, was the key treatment for patients experiencing symptomatic limitations due to obstructive HCM (oHCM) despite optimal medical therapy.^10–12^

Conventionally, the serial disease monitoring in patients with HCM has involved advanced cardiac imaging, including cardiac magnetic resonance imaging (CMR) and echocardiography, which can be limited by access and high costs.^13, 14^ Recently, artificial intelligence-enhanced electrocardiography (AI-ECG) has shown promise in tracking longitudinal changes linked to disease progression and treatment modification.^4, 15^ In a phase-2 clinical trial of oHCM, the AI-ECG phenotype for HCM improved on serial ECGs among patients receiving mavacamten.^4, 15^ However, there has not been a comprehensive assessment of the effects of SRT on the disease modification in the electrocardiographic signature of HCM. Moreover, despite prior advances, current deep learning-based approaches for evaluating disease modification have only been tested among highly selective patients enrolled in clinical trials.^4, 15^ These approaches have also relied on raw voltage signals that often not accessible to the end-user at the point-of-care.^4^

We leveraged an image-based deep learning strategy to evaluate disease modification by SRT for oHCM. For this, we deployed a model that detects HCM from images of 12-lead ECGs and evaluated ECGs from cohorts of individuals who underwent SRT at 3 distinct institutions. We used mavacamten recipients at one of these institutions as a positive control.

## METHODS

The Yale Institutional Review Board approved the study protocol and waived the need for informed consent as the study involves secondary analysis of pre-existing data. Patients who opted out of research studies at the Yale New Haven Health System (YNHHS) were not included.

### Data Sources

We included ECGs from patients who underwent SRT at three geographically distinct study sites: (1) Yale New Haven Health System (YNHHS), New Haven, CT, (2) Cleveland Clinic Foundation (CCF), Cleveland, OH, and (3) Atlantic Health System (AHS), Morristown, NJ **(Central Illustration)**.

The YNHHS represents the largest referral center in southern New England, comprising a network of 5 hospitals and a large outpatient medical group, serving areas of Connecticut, New York, and Rhode Island and providing care to over 2.7 million patients. CCF is the second largest private medical group practice in the USA, with 15 hospitals and over 200 outpatient locations, serving over 1 million patients annually in Northeast Ohio. The AHS is a large 7-hospital network, including the Morristown Medical Center, serving over 7.5 million patients in New Jersey. These study sites represent diverse healthcare systems, each with specialty HCM-care-clinic programs for patients with HCM and their family members.

### Study Population and Exposure

At YNHH, all patients who underwent SRT between 2013 and 2023 were included. For CCF and AHS, 100 and 145 patients with SRT were randomly selected from a list of SRT patients at each site and identified by the site-specific principal investigators (MD and MM, respectively). The ECGs of patients before and after SRT were identified using local ECG visualization software embedded with Epic EHR at each of the sites.

For patients who underwent SRT across sites, we identified the last ECG recorded before SRT and the most recent available ECG to identify the AI-ECG HCM probability before and after the procedures **(Central Illustration)**. At YNHHS, we also identified a corresponding procedural control group that included patients who underwent coronary artery bypass graft surgery (CABG) and surgical aortic valve replacement (SAVR). These were selected to identify the effects of cardiac surgical procedures on the HCM probability score among those without HCM. In this group too we included the closest ECGs recorded before the included procedure and the most recent ECG available after those procedures. Further, at YNHHS, we identified all ECGs from patients on treatment with mavacamten.

### Model Deployment

The study represents deployment of a previously developed AI-ECG model that detects HCM on ECG images.^1^ Briefly, the model was developed in the YNHHS using ECG images from patients with CMR-confirmed HCM who had not received mavacamten or undergone SRT, and validated in a demographically diverse cohort. This model had an area under the receiver operating characteristic curve of 0.95 (95% CI 0.93–0.97), with 92% sensitivity and 88% specificity for identifying patients with HCM.^1^ The model’s performance was comparable to other AI-ECG models for HCM that were developed for raw voltage data, but was able to detect HCM on ECG images, and is publicly available for research use at https://www.cards-lab.org/ecgvision-hcm.

At YNHHS, the model was deployed on ECG images with a voltage calibration of 10 mm/mV, with limbs and precordial leads arranged in four columns of 2.5-second each, representing leads I, II, and III; aVR, aVL, and aVF; V1, V2, and V3; and V4, V5, and V6. A full 10-second recording of the lead I signal represented the rhythm strip. These images were converted to greyscale and down-sampled to 300x300 pixels regardless of their original resolution using Python Image Library.^18^

For the external study sites, CCF and AHS, the model was applied to digital screenshots or smartphone photographs of ECG printouts, available as PNG images. These images followed a similar format with 4 columns, each with 2.5 seconds of each lead. However, the varied in the background hue (grey, orange, pink, or green), number of rhythm leads (1-3 leads), and the leads representing the rhythm leads (leads I, II, V1, or V5). Patient identifiers were cropped out or masked before the model application.

### Study Outcomes

For each ECG, the model output probability, or the AI-ECG HCM score was evaluated. This AI-ECG HCM score is the probability of the ECG being from a patient HCM, and represents the electrocardiographic HCM disease phenotype.^1, 15^ For each patient who underwent SRT across sites, we compared the AI-ECG HCM score for the closest ECG recorded before the SRT procedure with the latest available ECG. Similarly, we also calculated the changes in AI-ECG HCM scores among patients without HCM who underwent CABG or SAVR at YNHHS.

In a positive control experiment, we sought to evaluate the longitudinal effect of mavacamten on the AI-ECG HCM score. Among patients on mavacamten therapy, we assessed the AI-ECG HCM scores of these patients before and after the initiation of mavacamten. We compared the closest ECG recorded before the initiation of mavacamten therapy with the latest on-mavacamten ECG available. We also calculated the mean of all ECGs up to 3 years before therapy, representing the baseline ECG score, and longitudinally tracked the changes in the AI-ECG HCM score from the baseline, while patients were receiving mavacamten.

### Statistical Analyses

Categorical variables were reported as number (percentage, %), and continuous variables as mean (standard deviation [SD]) or median (interquartile range [IQR]), as appropriate. The AI-ECG HCM probabilities for each patient before and after the interventions were compared using the Wilcoxon rank sum test. In an exploratory analysis, we plotted a fourth-order generalized additive model curve for the change in AI-ECG HCM scores for on-mavacamten ECGs.

The statistical significance level was set at P < 0.05. All statistical analyses were executed using Python 3.11.2 and R version 4.2.0.

## RESULTS

### Study Population

A total of 70 patients underwent SRT at YNHHS, with a median age of 70 years (IQR 59-75). Of these patients, 32 (46%) were women, and 14 (20%) non-White **(Table 1)**. At YNHHS, 18 patients underwent alcohol septal ablation, and 52 underwent ventricular myectomy. At CCF, 100 patients underwent SRT, all ventricular myectomy, with a median age of 62 years (IQR 52-72), of whom 48 (48%) were women and 10 (10%) were non-White. The 145 patients who underwent SRT at AHS all were those who had undergone ventricular myectomy. This population had a median age of 60 years (IQR 48–67), 63 (43%) were women and 20 were non-White (13.8%) **(Table 1)**.

**Table 1.** Demographic Characteristics of the Study Population. **Abbreviations:** AHS, Atlantic Health System; CCF, Cleveland Clinic Foundation; YNHHS, Yale New Haven Health System.

| Characteristic |  | Septal Reduction Therapy |  |  | Mavacamten |
| --- | --- | --- | --- | --- | --- |
|  |  | YNHHS | CCF | AHS | YNHHS |
| Number |  | 70 | 100 | 145 | 36 |
| Age at SRT, Median [IQR] |  | 70 [59-75] | 62 [52-72] | 60 [48-67] | 65 [59-69] |
| Female Sex, N (%) |  | 32 (46%) | 48 (48%) | 63 (43%) | 15 (42%) |
| Race/Ethnicity,<br>N (%) | White | 56 (80%) | 90 (90%) | 125 (86%) | 28 (78%) |
|  | Black | 7 (10%) | 7 (7%) | 3 (2%) | 6 (17%) |
|  | Hispanic | 6 (8%) | - | 2 (1%) | 1 (3%) |
|  | Asian | - | 2 (2%) | 3 (2%) | - |
|  | Other | 1 (1%) | 1 (1%) | 8 (6%) | 2 (6%) |
|  | Missing | - | - | 4 (3%) | 3 (8%) |

In the procedural control population from YNHHS, representing all patients undergoing non-SRT major cardiovascular procedures at YNHHS, we included 4667 patients without HCM who underwent CABG (median age 72 years [64-79]; 1217 [27%] women; 1071 [21%] non-White) and 2316 who underwent SAVR (median age 73 years [64-81]; 711 [31%] women; 371 [15%] non-White), with ECGs recorded before and after the procedures **(Table S2)**.

At YNHHS, 36 patients receiving therapy with mavacamten were included, with a median age of 65 years (IQR 59-69), of whom 15 (42%) were women and 9 (25%) were non-White **(Table 1)**.

### Effect of SRT on AI-ECG HCM Phenotype

Among patients at YNHHS, the latest pre-SRT ECGs were recorded a median 28 days (IQR 7-64) before the procedure. The AI-ECG score before SRT was a median of 0.55 (IQR 0.24– 0.77). Following SRT, in the latest ECGs acquired a median 509 days (IQR 62-1156) after the procedure, there was no improvement in the HCM score (0.59 [IQR 0.40–0.75]; *p* = 0.43 by Wilcoxon signed-rank test) **(Table 2**, **Figure 1)**.

**Figure 1.**
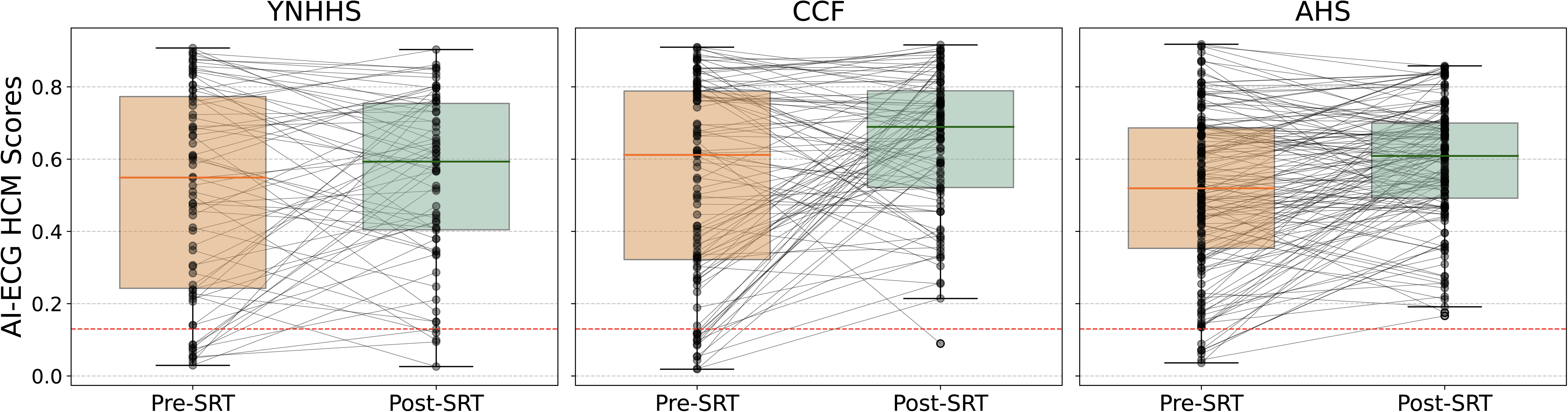
Changes in AI-ECG HCM score following septal reduction therapy. **Box and whisker plots for artificial intelligence-enabled electrocardiogram hypertrophic cardiomyopathy scores (AI-ECG HCM score) for images of closest electrocardiograms before and latest after septal reduction therapy.** Abbreviations: AHS, Atlantic Health System; AI, Artificial Intelligence; ECG, electrocardiogram; CCF, Cleveland Clinic Foundation; ECG, electrocardiogram; HCM, Hypertrophic Cardiomyopathy; SRT, Septal Reduction Therapy; YNHHS, Yale New Haven Health System.

**Table 2.** Artificial Intelligence-enabled Electrocardiogram Hypertrophic Cardiomyopathy Scores in Electrocardiograms Closest Before and Latest After Septal Reduction Therapy and Mavacamten Treatment. **Abbreviations:** AHS, Atlantic Health System; CCF, Cleveland Clinic Foundation; YNHHS, Yale New Haven Health System.

| **Intervention** | **Health System** | **Pre-Intervention**<br>**AI-ECG HCM**<br>**Score,**<br>**Median (IQR)** | **Post-**<br>**Intervention**<br>**AI-ECG HCM**<br>**Score,**<br>**Median (IQR)** | **P-value by**<br>**Wilcoxon**<br>**signed-rank**<br>**test** |
| --- | --- | --- | --- | --- |
| **Septal**<br>**Reduction**<br>**Therapy** | **YNHHS** | 0.55<br>(0.24-0.77) | 0.59<br>(0.40-0.75) | 0.428 |
|  | **CCF** | 0.61<br>(0.32-0.79) | 0.69<br>(0.52-0.79) | 0.003 |
|  | **AHS** | 0.52<br>(0.35-0.69) | 0.61<br>(0.49-0.70) | < 0.001 |
| **Mavacamten** | **YNHHS** | 0.41<br>(0.22–0.77) | 0.28<br>(0.11–0.50) | <0.001 |

At YNHHS, while the AI-ECG scores did not change in 18 patients who underwent alcohol septal ablation (0.58 [IQR 0.43–0.71] vs 0.41 [IQR 0.27–0.71], *p* = 0.08), the scores were higher post-SRT among 52 patients who underwent myectomy (0.51 [IQR 0.22–0.79] vs. 0.62 [IQR 0.44–0.76], *p* = 0.048) **(Table S1)**.

Among 100 patients at CCF, the AI-ECG HCM score increased post SRT (0.61 [IQR 0.32–0.79] vs 0.69 [IQR 0.52–0.79], *p* = 0.003). The AI-ECG score also increased at AHS from 0.52 (IQR 0.35–0.69) in the closest ECGs recorded a median of 45 days (IQR 5-119) before SRT to 0.61 (IQR 0.49–0.70; *p <* 0.001) acquired a median 664 days (IQR 113-1536) following the SRT procedure (**Figure 1**).

### Other Major Cardiovascular Procedures

In the procedural control analysis involving 4667 patients without HCM who underwent CABG at YNHHS, the median AI-ECG HCM scores changed from 0.13 (0.04-0.36) before the procedure to 0.19 (0.06-0.44) post CABG (*p <* 0.001) **(Table S3)**. Among 2316 patients who underwent SAVR, the AI-ECG HCM scores increased from 0.21 (0.05-0.48) to 0.24 (0.07-0.49) following the procedure (*p =* 0.01).

### Effect of mavacamten on AI-ECG HCM Phenotype

The median baseline AI-ECG HCM score before starting mavacamten, representing the mean of all ECGs recorded within 3 years before mavacamten initiation, was 0.46 (IQR 0.21–0.72). The median AI-ECG HCM score in the last ECG before starting mavacamten was 0.41 (0.22-0.77). At the end of a median follow-up period of 237 days (IQR 140–338), the median AI-ECG score decreased significantly to 0.28 (IQR 0.11–0.50, *p* <0.001) (**Table 2**, **Figure 2**).

**Figure 2.**
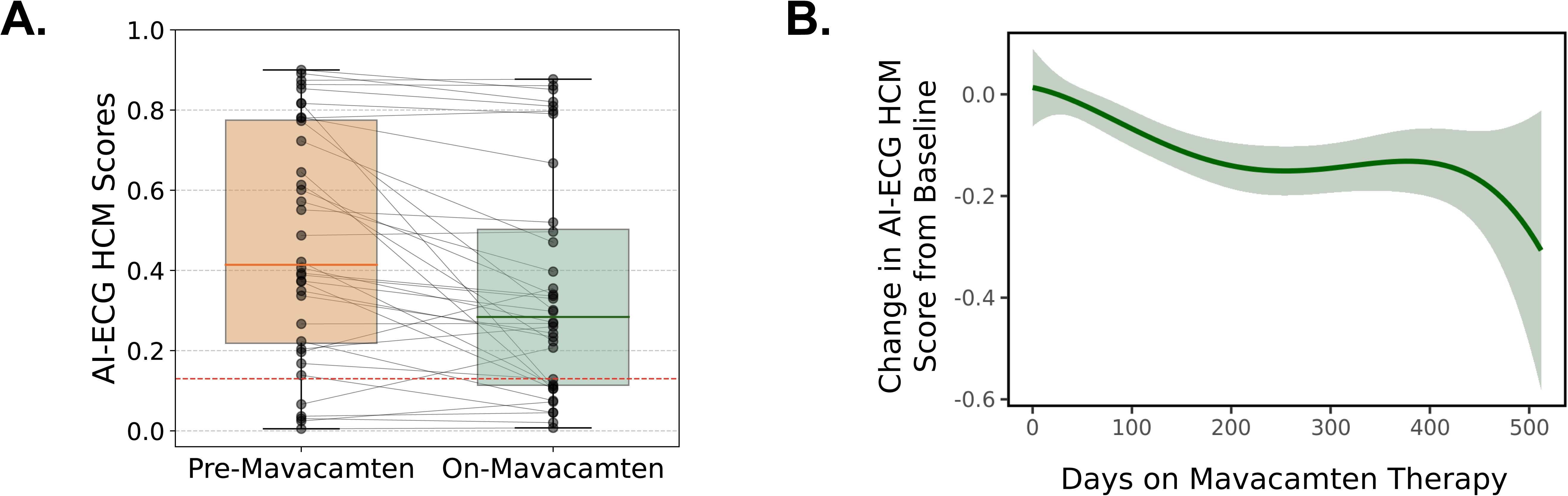
Changes in AI-ECG HCM score following mavacamten therapy. **(A) Box and whisker plot for artificial intelligence-enabled electrocardiogram hypertrophic cardiomyopathy scores (AI-ECG HCM score) for images of closest electrocardiograms before mavacamten initiation and latest on therapy (B) Fourth order generalized additive model curve for changes in AI-ECG HCM scores for on-mavacamten ECGs.** Abbreviations: AI, Artificial Intelligence; ECG, electrocardiogram; HCM, Hypertrophic Cardiomyopathy.

## DISCUSSION

In a study from 3 large medical centers, the electrocardiographic profile of HCM measured by an AI-ECG model deployed directly on images did not improve among patients who underwent SRT. In contrast, the AI-ECG HCM score significantly improved in a cohort of real-world HCM patients receiving mavacamten, replicating the effect observed in post-hoc analyses of a phase 2 randomized clinical trial of mavacamten.^4, 15^ The work highlights the differential effects of SRT and myosin inhibitors on the electrocardiographic marker of HCM while also defining a low-cost point-of-care strategy to track disease status directly on ECG images.

While the use of deep learning has been widely proposed for the detection of HCM from ECGs,^1–3^ the use of AI-ECG for tracking HCM substrate modification has been limited.^4, 15^ An evaluation of two signal-based HCM models applied to patients receiving mavacamten in a phase 2 clinical trial suggested an improvement in the electrocardiographic HCM phenotype on mavacamten therapy,^4, 15^ a finding consistent with our ECG-image-based assessment of a real-world cohort of patients on mavacamten at YNHHS. The contrast in the effects of SRT and cardiac myosin inhibition on AI-ECG HCM score may reflect different mechanisms for modification of the HCM substrate.^19, 20^ While SRT relieves obstructive physiology, an effect also observed with mavacamten, it does so by removing the anatomical obstruction without modifying the pathophysiology of diseased sarcomeres in other parts of the myocardium.^21^ In this respect, a significant decrease in the AI-ECG score among patients treated with mavacamten is concordant with its known systemic effects on myocardial remodeling and associated energetics.^8^

We observed a modest increase in the AI-ECG HCM scores following ventricular myectomy and other major cardiovascular surgical procedures, such as CABG and SAVR. These findings suggest that local post-surgical tissue modifications may modify the AI-ECG HCM score detected by the model. However, given the similar magnitude of increase across procedures, the AI-ECG HCM score does not suggest any improvement in the HCM score after ventricular myectomy. This is also substantiated by the lack of improvement in the AI-ECG HCM scores following alcohol septal ablation at YNHHS, representing the percutaneous approach for septal reduction.

A strength of our work is its multicentre design to evaluate the implications of AI-ECG to monitor disease. It also demonstrates the potential value and scalability of an image-based AI-ECG approach for disease monitoring in HCM and other disorders. The ECG image-based models for HCM can be used by health systems regardless of their underlying data format and also by individual clinicians who may only have access to ECGs as printouts or digital images.

### Study Limitations

Our study has several limitations. First, the number of patients receiving mavacamten therapy is low due to the relatively recent approval of the drug. Nonetheless, our study is the first to use ECG images to evaluate longitudinal changes in the AI-ECG HCM phenotype in a real-world patient care setting. Moreover, our findings are consistent with the effect of mavacamten observed in previous reports that used raw ECG signal data from a phase-2 clinical trial.^4, 15^ Second, the AI-ECG phenotype among patients with HCM may change longitudinally over the course of disease, with early stages of disease potentially presenting with a lower pathophysiological ECG signature. However, we included the closest ECG recorded before the SRT procedures across sites to capture the advanced stages of disease requiring surgical intervention. Third, there may be acute local inflammation or edema immediately following major cardiovascular procedures, which could potentially alter the AI-ECG phenotype. However, we used the most recent available ECG for each patient who underwent SRT, with the SRT to post-SRT ECG separated by a median 509 days to ensure that the AI-ECG HCM score was not merely reflective of acute postoperative changes.

## CONCLUSIONS

In conclusion, our study demonstrates a measurable difference in the myocardial effects of SRT and cardiac myosin inhibition detectable on ECG images and suggests a potential strategy for serial monitoring of pathophysiological improvement in HCM at the point-of-care.

## CLINICAL PERSPECTIVES

### Core Clinical Competencies

The recent approval of mavacamten for management of obstructive hypertrophic cardiomyopathy (HCM) offers an emerging oral alternative to the surgical or percutaneous reduction of the interventricular septum (SRT), which has previously been the standard of treatment. Our use of artificial intelligence on electrocardiographic (ECG) images demonstrates the contrasting physiological effects of the procedural and pharmacological therapy. In patients with obstructive HCM across 3 geographically distinct US sites, septal reduction therapy did not improve the ECG phenotype of HCM, which contrasts with cardiac myosin inhibition that resulted in significant improvements.

### Translational Outlook Implications

The use of artificial intelligence-enhanced ECG presents a key opportunity for feasibly monitoring the response to treatment in HCM, which has previously relied on advanced cardiac imaging such as echocardiography and cardiac magnetic resonance imaging. Our image-based strategy suggests a practical and scalable strategy for the assessment of the HCM phenotype at the point-of-care.

## ABBREVIATIONS

AI: Artificial Intelligence
AHS: Atlantic Health System
AUROC: Area Under the Receiver Operating Characteristic Curve
CABG: Coronary Artery Bypass Graft Surgery
CCF: Cleveland Clinic Foundation
CMR: Cardiac Magnetic Resonance Imaging
DPI: Dots Per Inch
ECG: Electrocardiograms
HCM: Hypertrophic Cardiomyopathy
IQR: Interquartile Range
oHCM: Obstructive Hypertrophic Cardiomyopathy
SAVR: Surgical Aortic Valve Replacement
SRT: Septal Reduction Therapy
YNHHS: Yale New Haven Health System

## Data Availability

The dataset cannot be made publicly available because they are electronic health records. Sharing this data externally without proper consent could compromise patient privacy and would violate the Institutional Review Board's approval for the study. The code for the study is available from the authors upon request.

---

**Central Illustration.**
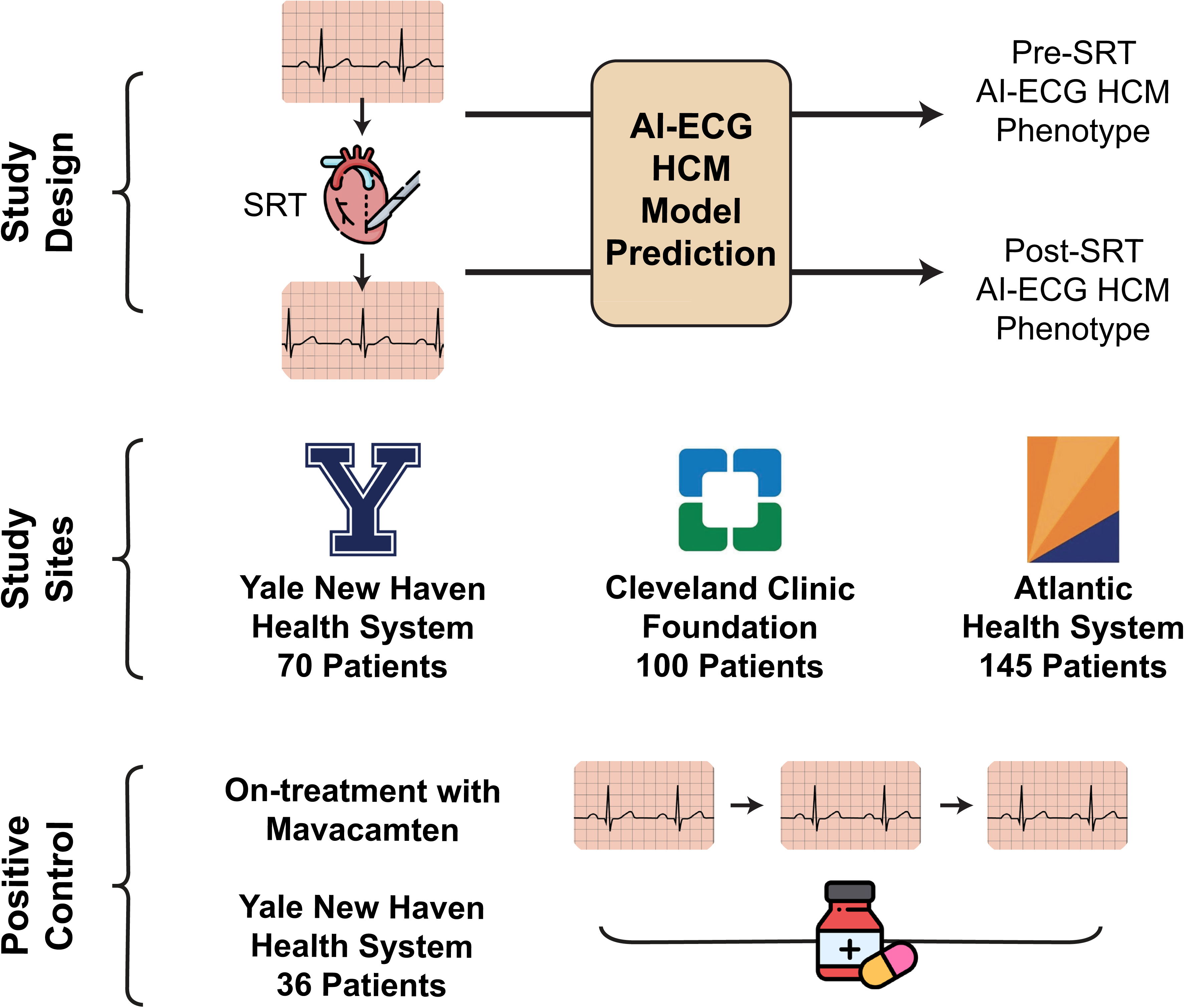
Title: Study Design and Population. **Caption: A Multicenter Evaluation of Changes in the Artificial Intelligence-enabled Electrocardiogram Hypertrophic Cardiomyopathy Scores following Septal Reduction Therapy and Mavacamten Treatment.** Abbreviations: AI, Artificial Intelligence; ECG, electrocardiogram; HCM, Hypertrophic Cardiomyopathy.

