## Supplementary material for "A Multicenter Evaluation of the Impact of Therapies on Deep Learning-based Electrocardiographic Hypertrophic Cardiomyopathy Markers": Online Supplement

**Supplementary Table S1. Artificial Intelligence-enabled Electrocardiogram Hypertrophic Cardiomyopathy Scores Before and After Septal Reduction Therapy at Yale New Haven Hospital. Abbreviations:** AI, Artificial Intelligence; ECG, Electrocardiogram; HCM, Hypertrophic Cardiomyopathy; IQR, Interquartile Range.

| **Intervention** | **Number of Patients** | **Pre-Intervention AI-ECG HCM Score,**  **Median (IQR)** | **Post-Intervention**  **AI-ECG HCM Score,**  **Median (IQR)** | **P-value by Wilcoxon signed-rank test** |
| --- | --- | --- | --- | --- |
| **Alcohol Septal Ablation** | 18 | 0.58 (0.43-0.71) | 0.41 (0.27-0.71) | 0.081 |
| **Ventricular Myectomy** | 52 | 0.51 (0.22-0.79) | 0.62 (0.44-0.76) | 0.048 |

**Supplementary Table S2. Demographic Characteristics of the Patients undergoing Coronary Artery Bypass Graft Surgery and Surgical Aortic Valve Replacement at Yale New Haven Health System.** **Abbreviations:** CABG, Coronary Artery Bypass Graft Surgery; IQR, Interquartile Range; SAVR, Surgical Aortic Valve Replacement.

| **Characteristic** | | **Patients Undergoing**  **CABG** | **Patients Undergoing SAVR** |
| --- | --- | --- | --- |
| **Number** | | 4667 | 2316 |
| **Age at Procedure, Median [IQR]** | | 72 [64-79] | 73 [64-81] |
| **Female Sex, N (%)** | | 1217 (27%) | 711 (31%) |
| **Race/Ethnicity,**  **N (%)** | **White** | 3596 (79%) | 1945 (85%) |
|  | **Black** | 349 (8%) | 136 (6%) |
|  | **Hispanic** | 386 (8%) | 117 (5%) |
|  | **Asian** | 104 (2%) | 25 (1%) |
|  | **Other** | 78 (2%) | 29 (1%) |
|  | **Missing** | 67 (1%) | 32 (1%) |

**Supplementary Table S3. Artificial Intelligence-enabled Electrocardiogram Hypertrophic Cardiomyopathy Scores Before and After Coronary Artery Bypass Graft Surgery and Surgical Aortic Valve Replacement. Abbreviations:** AI, Artificial Intelligence; ECG, Electrocardiogram; HCM, Hypertrophic Cardiomyopathy; IQR, Interquartile Range.

| **Intervention** | **Number of Patients** | **Pre-Intervention AI-ECG HCM Score,**  **Median (IQR)** | **Post-Intervention**  **AI-ECG HCM Score,**  **Median (IQR)** | **P-value by Wilcoxon signed-rank test** |
| --- | --- | --- | --- | --- |
| **Coronary Artery Bypass Graft Surgery** | 4667 | 0.13 (0.04-0.36) | 0.19 (0.06-0.44) | < 0.001 |
| **Surgical Aortic Valve Replacement** | 2316 | 0.21 (0.05-0.48) | 0.24 (0.07-0.49) | 0.014 |
